# Derived Intoxicating Cannabis Vape Product Attributes and Marketing in an Online Retail Environment

**DOI:** 10.1101/2025.01.22.25320970

**Authors:** Julia Chen-Sankey, Cassidy R. LoParco, Kathryn La Capria, Siyan Meng, Rosanna Mazzeo, Neha Vijayakumar, Amanda Y. Kong, Kayla K. Tillett, Carla Berg, Matthew E. Rossheim

**Affiliations:** Rutgers School of Public Health, Piscataway, NJ; Rutgers Institute for Nicotine & Tobacco Studies, New Brunswick, NJ; Milken Institute School of Public Health, George Washington University, Washington, DC; Wake Forest University School of Medicine, Winston-Salem, NC; College of Public Health, University of North Texas Health Science Center, Fort Worth, TX; George Washington Cancer Center, George Washington University, Washington, DC

**Author notes:** **Corresponding author:** Julia Chen-Sankey, PhD, MPP, Rutgers School of Public Health, 303 George St. Room 525, New Brunswick, NJ 08901. **Disclosure:** Dr. Kong has served as a paid expert consultant in litigation against the tobacco industry.

**Keywords:** Cannabis marketing, cannabis advertising, derived cannabis vape products, online retailers, cannabis product descriptions

## Abstract

**Introduction:** The 2018 Farm Bill unintentionally allowed the proliferation of derived intoxicating cannabis vape products (DICVPs), raising concerns about associated health risks. To inform public health prevention efforts, this study analyzed the product attributes and marketing features of DICVPs in an online retail environment.

**Methods:** In 2023, we extracted information on product attributes and descriptions of 490 DICVPs from two online retail websites with high web traffic. In 2024, two trained coders thematically coded product descriptions for their product characteristics and marketing features.

**Results:** Overall, 95 unique brands and 26 unique intoxicating cannabinoids were identified. The most frequent marketing features were overall vape product design and use (99.0%), including vaping satisfaction, discreetness, convenience, and use instructions. Regulation and compliance messages (91.6%) were also prevalent, including lab testing for additives and/or chemicals, health warnings, hemp-derived labels, references to the 2018 Farm Bill, and FDA approval statements. Other prominent themes included: flavor and sensation claims (79.6%, i.e., flavor variety, fruit flavors); psychoactive effect claims (43.3%, e.g., potency or expected user experience); product quality claims (38.4%, e.g., “quality,” “natural,” “purity”); and other positive effect claims (33.9%, e.g., mood enhancement, relaxation).

**Discussion:** The DICVP online marketplace is highly fragmented with a variety of brands and intoxicating compounds. Common marketing strategies promoting appealing flavors and positive vaping experiences may increase product use interest among young people. Features related to product legality and quality may reduce perceived barriers and risks of using products. Continuous monitoring of the DICVP marketplace is needed to inform policymaking.

## INTRODUCTION

Since the passage of the 2018 Farm Bill (the Agriculture Improvement Act), which excluded hemp (≤0.3% delta-9 tetrahydrocannabinol [THC]) from the U.S.’s list of controlled substances (1), many new types of derived intoxicating cannabis products (DICPs) are being sold widely throughout the U.S. market (e.g., delta-8 THC, delta-10 THC) (2–6).

Studies have found that DICPs have proliferated across the country and come in various forms, the most common of which are edibles (e.g., gummies, desserts, beverages) and vape products (e.g., cartridges, pods, or disposables) (2–5). DICPs are widely sold in gas stations, convenience stores, smoke/vape/head shops, and online, often in ways that are both accessible and attractive to young people (2–4,7,8). Consistent with concerns regarding youth access and appeal, national data from 2023 shows that one in nine 12th graders (11.4%) reported using delta-8 THC in the past year (9). This is likely an underestimation of DICP use, considering the dozens of other DICPs beyond delta-8 THC (2–4,10).

Similar to traditional cannabis products, DICPs may result in acute health effects, including anxiety, lethargy, and impaired coordination (11). Regular and long-term use of DICPs, just like traditional cannabis, may lead to cannabis dependence as well as psychosis and other mental health illnesses (11). Use of DICPs also carries its own risks unique to those of traditional cannabis. Due to their largely unregulated status, DICPs frequently feature misleading or inaccurate labeling and are often not tested for potency or contamination (12,13). Reports indicate some have been tainted with heavy metals, and they have been linked to thousands of incidents of accidental ingestion, some of which required medical attention, especially among young individuals (11). At the same time, the availability and use of higher potency cannabis products, including DICPs, have been on the rise, which is concerning because high-potency cannabis products and younger age are both primary risk factors for developing cannabis dependence and cannabis-induced psychosis (14). Therefore, stricter DICP-related regulations and restrictions are needed to protect public health.

Derived intoxicating cannabis vape products (DICVPs) are a commonly sold form of DICPs in brick-and-mortar and online stores (2–5). DICVPs resemble the appearance of traditional nicotine vape products and may be especially attractive to young people due to the highly convenient and discreet nature of product use. In addition, almost all DICVPs sold in the marketplace come in fruit and candy flavors, which research indicates is one of the most appealing product attributes for using nicotine vape products among youth and young adults (15– 17). Evidence also shows that the potency of cannabis concentrates inhaled through vaping is substantially higher than for flower cannabis products (18,19), potentially increasing the risk of dependence or negative effects. Cannabis vaping also has a more rapid onset than other products (e.g., edibles, capsules), which might cause users to experience immediate adverse and psychoactive effects (20).

There is a lack of systematic understanding of the product attributes and marketing strategies used by the industry to promote DICVPs. Investigating product attributes is crucial because these attributes—such as potency, price, and type of intoxicating cannabinoid(s)—can significantly impact perceptions, behavior, and health outcomes (21,22). Examining the marketing features—such as flavor, product design, risk, and benefit claims—of these products can shed light on their potential appeal to young people (15–17). Marketing features such as appealing flavors, sleek and discreet device design, reduced harm claims, and social and mood enhancement benefits can make DICVPs seem to be more attractive and healthier to use for young consumers (23). Examining the presence of regulation-related claims may further provide insights into how consumers may perceive the legality, safety, and risks of using the products.

The current study used information from two highly visited DICP retail websites to describe product attributes and marketing features of DICVPs (2). Online retail was particularly important for this investigation due to the inadequate age verification practices for selling these products to minors and the wide availability of DICVP marketing and sales throughout the country (24,25).

## METHODS

### Data Source and Measures

We recorded information on product attributes and descriptions of the DICVPs sold on two prominent U.S.-based cannabis websites, *Element Vape* (elementvape.com) and *Premium Delta-8 and CBD* (delta8resellers.com) between May and August 2023. In April 2023, both websites had a high volume of web traffic, with 2.1 million visits to *Element Vape* and 547,500 visits to *Premium Delta-8 and CBD* (2). Detailed information about why the two websites were chosen and the procedure of recording the information is documented elsewhere (2). Specifically, we recorded the information related to the product attributes of each DICVP (*n*=490), including brand name, vape product modality, pack size, price, and the type(s) of intoxicating cannabinoid(s), such as Delta-8 THC. We also recorded product descriptions for each DICVP.

### Coding Procedure

In June-August 2024, we conducted a content analysis of the information recorded to capture product attributes and descriptions of the DICVPs from the two websites using a deductive and inductive approach. The deductive approach involved leveraging existing research on cannabis product marketing features and empirical evidence of outcome expectancies for using cannabis and vape products (13,26,27). The inductive approach involved conducting a qualitative review of the product descriptions to develop and generate additional codes related to DICVP marketing features. Codes were not mutually exclusive. Since all DICVPs examined in the study were flavored, we did not include flavor-related codes to describe product attributes.

However, we did generate a series of flavor-related codes for marketing features. Three trained coders from the research team first reviewed the codebook and tested the codes by independently coding a sample of the product descriptions for marketing features. The research team then met to discuss and confirm the final codes. Inter-rater reliability was established through two rounds of double coding using a sub-sample (*n*=50 each round) of product descriptions. Discrepancies were resolved and the codebook was refined through discussion during team meetings. After the second round of double-coding, we reached an average inter-rater reliability of Krippendorff’s alpha >0.8 across codes (28). Discrepancies in coding were further discussed to improve reliability. The remaining product descriptions were then randomly split between coders for content coding. The final codebook of marketing features consisted of six categories covering 29 codes (see **Table 3** for all marketing feature codes and definitions).

### Data Analysis

Descriptive statistics (percent, mean, and standard deviation [SD]) were generated using Stata v.18. Institutional Review Board approval was not required because this study utilized publicly available data and was not considered human subject research under 45 CFR 46.102(d).

## RESULTS

### Product Attributes

The 490 DICVPs (29.2% cartridge and 70.8% disposable) were from 95 unique brands (**Table 1**). The 10 most common brands were Delta Extrax (6.1%), Cake (5.9%), Looper (4.5%), Honeyroot (4.1%), Mellow Fellow (3.9%), Binoid (3.9%), TRE House (3.3%), Modus/Medusa (3.1%), Exhale (2.9%), and URB (2.9%).

**Table 1.** Brands (n=95) of derived intoxicating cannabis vape products (n=490)

| **Brand** | **%** | **n** |
| --- | --- | --- |
| Delta Extrax | 6.1 | 30 |
| Cake | 5.9 | 29 |
| Looper | 4.5 | 22 |
| Honeyroot | 4.1 | 20 |
| Binoid | 3.9 | 19 |
| Mellow Fellow | 3.9 | 19 |
| TRE House | 3.3 | 16 |
| Modus/Medusa | 3.1 | 15 |
| Exhale | 2.9 | 14 |
| URB | 2.9 | 14 |
| Purlyf | 2.7 | 13 |
| Flying Monkey | 2.5 | 12 |
| Kalibloom | 2.5 | 12 |
| ELYXR | 2.2 | 11 |
| Pacha | 2.2 | 11 |
| HiXotic | 2.0 | 10 |
| ZAZA | 1.8 | 9 |
| Ghost | 1.6 | 8 |
| Torch | 1.6 | 8 |
| Maui Labs | 1.4 | 7 |
| Trippy Sugar | 1.4 | 7 |
| ASTRO EIGHT | 1.2 | 6 |
| Dimo | 1.2 | 6 |
| Geek'd | 1.2 | 6 |
| Hidden Hills | 1.2 | 6 |
| Kik | 1.2 | 6 |
| Wild Orchard | 1.2 | 6 |
| Exodus | 1.0 | 5 |
| Happi | 1.0 | 5 |
| Ocho Extracts | 1.0 | 5 |
| Space Gods | 1.0 | 5 |
| D8Co. | 0.8 | 4 |
| Frozen Fields | 0.8 | 4 |
| Koi | 0.8 | 4 |
| Packwoods | 0.8 | 4 |
| Sugar | 0.8 | 4 |
| Zombi | 0.8 | 4 |
| Baked | 0.6 | 3 |
| Cali Reserve | 0.6 | 3 |
| ELF THC | 0.6 | 3 |
| Fume | 0.6 | 3 |
| Galaxy Treats | 0.6 | 3 |
| Ript | 0.6 | 3 |
| STIIIZY | 0.6 | 3 |
| STNR | 0.6 | 3 |
| Treetop | 0.6 | 3 |
| 3Chi | 0.4 | 2 |
| Blazed | 0.4 | 2 |
| Canna River | 0.4 | 2 |
| Chapo Extrax | 0.4 | 2 |
| Crooked Creations | 0.4 | 2 |
| Delta Man | 0.4 | 2 |
| Dozo | 0.4 | 2 |
| Esco Bars | 0.4 | 2 |
| Goo'd | 0.4 | 2 |
| HIGH | 0.4 | 2 |
| Half Bak'd | 0.4 | 2 |
| Hazy Extrax | 0.4 | 2 |
| Juicy Kush | 0.4 | 2 |
| Just | 0.4 | 2 |
| Kream | 0.4 | 2 |
| Kush Burst | 0.4 | 2 |
| LITTO | 0.4 | 2 |
| Punch Labs | 0.4 | 2 |
| STILO | 0.4 | 2 |
| Slap Hemp | 0.4 | 2 |
| Space Monkey | 0.4 | 2 |
| Stoney | 0.4 | 2 |
| Strange Clouds | 0.4 | 2 |
| Tiny Dancer | 0.4 | 2 |
| Trip Drip | 0.4 | 2 |
| Two Hawk | 0.4 | 2 |
| Tyson | 0.4 | 2 |
| Wonderbrett | 0.4 | 2 |
| 10X | 0.2 | 1 |
| 8THS | 0.2 | 1 |
| Ace bar | 0.2 | 1 |
| Alibi | 0.2 | 1 |
| Artisan | 0.2 | 1 |
| Bad Days | 0.2 | 1 |
| Cali Extrax | 0.2 | 1 |
| Carats | 0.2 | 1 |
| Delta Munchies | 0.2 | 1 |
| Eighty Six | 0.2 | 1 |
| Fresh | 0.2 | 1 |
| High Nightcap | 0.2 | 1 |
| High Times | 0.2 | 1 |
| Hyper | 0.2 | 1 |
| Kush Kolectiv | 0.2 | 1 |
| Kynn | 0.2 | 1 |
| Moon Wlkr | 0.2 | 1 |
| Pax Era | 0.2 | 1 |
| Puff | 0.2 | 1 |
| Space Walker | 0.2 | 1 |
| Wavy | 0.2 | 1 |

There were 26 different intoxicating cannabis compounds contained within these 490 DICVPs (**Table 2**). The most common compound was Delta-8 THC (67.8%), followed by THC-P (39.0%), HHC (20.2%), Delta-10 THC (16.3%), THC-A (13.9%), Delta-9 THC (12.0%), THC-H (11.2%), THC-B (10.2%), THC-JD (9.0%), and THC-X (7.1%). Most of the products (62.9%) were blends, containing two or more intoxicating cannabinoids. The average number of intoxicating cannabinoids in each DICVP was 2.4 (range 1 to 8, SD = 1.5).

**Table 2.** Types of cannabinoids (n=26) in derived intoxicating cannabis vape products (n=490)

| **Cannabinoid type** | **Derived intoxicating cannabis vape products** |  |
| --- | --- | --- |
|  | **%** | **n** |
| Delta-8 THC | 67.8 | 332 |
| THC-P | 39.0 | 191 |
| HHC | 20.2 | 99 |
| Delta-10 THC | 16.3 | 80 |
| THC-A | 13.9 | 68 |
| Delta-9 THC | 12.0 | 59 |
| THC-H | 11.2 | 55 |
| THC-B | 10.2 | 50 |
| THC-JD | 9.0 | 44 |
| THC-X | 7.1 | 35 |
| HHC-P | 6.5 | 32 |
| Delta-11 THC | 5.9 | 29 |
| PHC | 5.9 | 29 |
| THC-O | 3.1 | 15 |
| THC-m | 2.9 | 14 |
| THC-V | 2.7 | 13 |
| Delta-6 THC | 2.5 | 12 |
| HHC-O | 2.0 | 10 |
| HXC | 1.0 | 5 |
| HXC-P | 0.8 | 4 |
| THC-PO | 0.6 | 3 |
| Delta-9O THC | 0.4 | 2 |
| HHC-R | 0.4 | 2 |
| THD | 0.4 | 2 |
| HCP | 0.2 | 1 |
| HXCO | 0.2 | 1 |

Most DICVPs were offered in a pack size of one (94.9%), and few came in packs of two (4.5%) or three (0.6%). Retail prices for the DICVPs ranged from $7.99 to $69.95, with an average price of $24.22 (SD = $8.10). The average price was $22.2 for cartridges (SD = $9.9) and $26.5 for disposables (SD = $7.9).

### Product Marketing Features

**Table 3** presents the frequencies of product marketing features in the DICVP descriptions, along with the definitions and examples. The most common marketing feature was vape product design and use (99.0%), defined as characteristics or features of product design and any aspects related to the user vaping experience. This included product design (97.6%, e.g., product color, shape), vaping satisfaction (28.2%, e.g., “leaving you satisfied after each hit”), discreetness/convenience (28.0%, e.g., “discreet,” “easy to use,” “on the go”), and use instructions (12.2%, e.g., how to power up the product).

**Table 3.** Product marketing feature categories and codes of the 490 online derived intoxicating cannabis vape products.

| Marketing Feature Categories | Definition | % | n | Example |
| --- | --- | --- | --- | --- |
| **Vape Product Design and Use** |  | **99.0** | **485** |  |
| Product design | Mentions any product design aspects, such as product colors, shapes, or specific features (i.e., heating elements, stainless steel hardware) or type of vape product (cartridge or disposable). | 97.6 | 478 | The Disposables are USB-C rechargeable and feature a slim and sleek design. |
| Vaping satisfaction | Mentions the user will experience satisfaction from using the product and may include phrases like “enjoy the experience,” “satisfying experience,” “positive vaping sensation.” | 28.2 | 138 | Leave you satisfied after each hit. |
| Discreetness/convenience | Includes claims that the product is convenient, discreet, easy to use, and use on the go. | 28.0 | 137 | Its sleek and compact design ensures easy portability, allowing you to enjoy your favorite strains anytime, anywhere. |
| Use instructions | Includes general instructions on how to use the products (e.g., how to power up the product). | 12.2 | 60 | To switch the device on or off, simply click the power button 5 times. Activate the pre-heat mode by clicking the power button 2 times, and once the light switches off, press and hold the power button to start vaping. |
| **Regulation and Compliance** |  | **91.6** | **449** |  |
| Lab tested for additives and chemicals | References the product being lab-tested or third-party tested for addictive and chemicals or free of chemicals, bacteria, additives, fillers, or other harmful substances etc. | 62.5 | 306 | Tested by third-party labs, the Delta-8 disposables are free of any impurities, pesticides, and toxins. |
| Health warning | Includes the California state warning about using cannabis products. | 51.2 | 251 | **WARNINGS:** Consuming this product can expose you to chemicals including beta-myrcene, which is known to the State of California to cause cancer and Δ9-Tetrahydrocannabinol, which is known to the State of California to cause birth defects or other reproductive harms. |
| Hemp-derived label | Includes claims about the product being hemp-derived or made of hemp. | 45.3 | 222 | Made from USA grown hemp. |
| 2018 Farm Bill | Includes claims about the product being Farm Bill compliant or federally legal under the Farm Bill. | 32.7 | 160 | These cartridges are 2018 farm bill compliant containing less than 0.3% delta-9 THC. |
| FDA approval statement | Includes the statement that the product has not been evaluated or approved by the FDA. | 6.3 | 31 | These statements have not been evaluated by the Food and Drug Administration. |
| **Flavor and Sensation** |  | **78.6** | **390** |  |
| Flavor variety | Includes claims that the product comes in a variety of flavor options. | 62.9 | 308 | Get ready to unleash the flavor explosion with a variety of mouthwatering options! |
| Fruit flavor | Describes the product to taste like fruit(s) (e.g., citrus flavors, strawberry, watermelon, grape, etc.). | 38.2 | 187 | Blackberry: The juicy sweetness of blackberries can now be enjoyed with the uplifting effects of Delta-8 THC! |
| General flavor description | Includes terms used to describe the product’s flavor (big flavor, flavorful, etc.) or general sensations of the product’s flavor (tasty, delicious, refreshing, etc.). | 30.2 | 148 | Try our tasty and satisfying Delta-8 Live Resin Disposables for delicious flavors and a powerful pinch of euphoria! |
| Other flavor | Includes the product’s flavor name that is not a fruit or words | 29.4 | 144 | Each hit delivers a savory blast of sweet and |
|  | that describe the product to taste like non-fruit related flavors such as mint, floral, dessert, candy (cake, mint, candy, etc.). |  |  | creamy vanilla undercut with hints of earthy pepper and bready sourness. |
| Flavor sensations | Includes terms to describe the tastes, smells, and textures that make up the product's flavor, such as "sweet," "sour," and "spicy." | 28.2 | 138 | Dominating the terpene profile is the subtle tartness of fresh berries that give way to a sweet aftertaste that saturates your palate. |
| **Psychoactive Effects** |  | **43.3** | **212** |  |
| Potency descriptors | Includes product potency such as powerful, potent, or strong. | 35.3 | 173 | Each disposable device contains 1.8 grams of the most potent distillate available. |
| Psychoactive effects | Includes the psychoactive effects of the product on the user such as feeling high, stoned, or buzzed. | 14.7 | 72 | Disposables provides an uplifting and functional buzz like no other. |
| Psychoactive effect use instructions | Includes a statement about how the product should be vaped, the number of puffs that one should be starting with, and the number of minutes for waiting for adjusting for tolerance. | 9.0 | 44 | We suggest starting with 1 puff and hold each hit 5-10 seconds and wait 30 mins to test your tolerance. |
| **Product Quality** |  | **38.4** | **188** |  |
| Natural | Includes the word 'natural.' | 16.5 | 81 | Flavored with all-natural terpenes. |
| Quality | Includes the word 'quality.' | 12.5 | 43 | The most potent and highest quality distillate available. |
| Purity | Includes the word 'pure/purity/purified.' | 10.6 | 52 | 1 gram of delta-8 cannabinoids harvested from legal hemp plants, tested to ensure potency and purity. |
| Premium | Includes the word 'premium.' | 10.2 | 50 | The Delta-9 disposable vape pen contains 3 grams of premium hemp-derived Delta 9 and Delta 8 distillate. |
| Organic | Includes the word 'organic.' | 8.8 | 43 | All disposables are derived from USA Grown Hemp with high end organic terpenes. |
| **Other Positive Effects** |  | **33.9** | **166** |  |
| Promoting positive mood or feelings | Includes claims that product use is associated with positive mood or feelings such as happiness and euphoria or associated with avoiding negative mood such as feeling down. | 24.5 | 120 | 2 grams of delta-8 THC packed into a convenient 510 thread cartridge to deliver elevated happiness. |
| Relaxation/tension reduction | Includes claims that product use is associated with relaxation or tension reduction and may include terms such as relaxed, calm, or chill. | 19.6 | 96 | User feedback describes effects of these disposable vapes as relaxing and perfect for unwinding after a long day. |
| Focus/creativity | Includes claims that product use will result in better focus or concentration, alertness, creativity, awareness, etc. | 5.3 | 26 | Elevate your senses, unlock your creativity, and embrace the possibilities with these 2-grams disposables. |
| Health effects | Includes claims that product use is associated with physical health benefits (e.g., pain, cancer), mental health benefits (e.g., stress, anxiety), general therapeutic health benefits, or sleep benefits. | 4.1 | 20 | Dealing with feelings of anxiety or physical discomfort may appear more manageable too. |
| Socialization effects | Includes claims that product use may help with friendship, familiarity, romance, and closeness with others. | 2.2 | 11 | Perfect for daytime use, this all-in-one vape will obliterate creative roadblocks and is great for social gatherings thanks to its talkative effects. |

Regulation and compliance, defined as warning statements or claims about the product being compliant with cannabis laws or regulations, appeared in 91.6% of the descriptions. Specifically, this included messages that the product was lab tested for additives and/or chemicals (62.5%, e.g., “tested by third-party labs, this product is free of impurities and toxins”), health warnings (51.2%, the California state warning about health effects from using cannabis products), the claim that the product was derived from hemp (45.3%, “made from USA grown hemp”), the 2018 Farm Bill (32.7%, “these cartridges are 2018 Farm Bill compliant, containing less than 0.3% delta-9 THC”), and non-FDA approval statements (6.3%, “these statements have not been evaluated by the FDA”). During coding, we found that all identified health warnings were from California and specific to Delta-9 THC products.

Flavor and sensation claims, defined as specific flavor names and general flavor or flavor sensation references describing the taste of the products, were noted in 79.6% of product descriptions. This included flavor variety (62.9%, e.g., claim that the product comes in a variety of flavors), fruit flavors (38.2%, e.g., “strawberry,” “watermelon”), general flavor descriptions (30.2%, “big flavor,” “delicious”), other flavor (29.4%, flavor that does not fall under fruit flavor categories, e.g., “mint,” “cake,” “cookies”), and flavor sensations (28.2%, terms that describe the tastes, smells, and textures of a flavor, e.g., “sweet,” “spicy”). It is worth noting that although we did not code product strains, some strain names may overlap with flavor names, such as “wedding cake” and “gelato.” Many flavors with trademark names of desserts and snacks were also identified during coding, including “Fruity Pebbles,” “Gushers,” “Hubba Bubba,” “Thin Mint Cookies,” and “Double Bubble.”

Psychoactive effects (43.3%) were defined as descriptors, claims, or instructions related to the psychoactive properties or effects of a product. Specifically, this included product potency descriptors (35.3%, keywords such as “powerful,” “potent,” and “strong”), psychoactive effects on the user (14.7%, e.g., feeling high, buzzed, or stoned), and user instructions on managing psychoactive effects (9.0%, e.g., the number of puffs that one should be starting with). Product quality claims (38.4%) highlighted the products’ ingredients or standards that may communicate low risks of harm to consumers. These claims commonly included the keywords “natural” (16.5%, e.g., “flavored with all-natural terpenes”), “quality” (12.5%, e.g., “the highest quality distillate available”), “purity” (10.6%, e.g., “highly potent”), “premium” (10.2%, e.g., “contains 3 grams of premium hemp-derived Delta 8 distillate”), and “organic” (8.8%, e.g., “has high-end organic terpenes”). Lastly, other effects from product use (33.9%) included content that promoted positive mood and feelings (24.5%, e.g., happiness, euphoria), relaxation and tension reduction (19.6%, e.g., relaxed, chill, calming effect), focus/creativity (5.3%, e.g., better focus, concentration, creativity), positive health effects (4.1%, physical health benefits [e.g., pain, cancer], mental health benefits [e.g., anxiety, depression, sleep benefits], and socialization effects (2.2%, e.g., closeness with others, improves friendships).

## DISCUSSION

This is one of the first studies to systematically examine the product attributes and marketing features of a large sample of DICVPs sold by online retailers with a high volume of web traffic. Similar to the overall DICP marketplace (2), the market of DICVPs is highly fragmented, as we documented more than 95 brands and 490 products being marketed and sold on these two websites. The most common brand, Delta Extrax, only comprised 6% of the products included in the study. It is also worth noting that more than 60% of the examined DICVPs are Delta-8 THC, for which youth use prevalence and negative health consequences have increased in recent years (9,29,30). The variety of types of intoxicating cannabinoids also signifies the diversity of the marketplace. The fact that most use a blend of cannabinoids may further add challenges to understanding the potential health implications of use. This fragmented market concentrated with small brands and diverse products may increase challenges for regulatory and enforcement efforts and may overwhelm and confuse consumers, potentially leading to product misuse or health risks.

We found that the most frequently promoted marketing features were those describing overall vape product design and use that were not specific to cannabis products. This information mostly included appealing aspects of vape products, which highlighted the “fun, coolness, and high technology” aspects of the product, as well as the “satisfying vaping experience” and the “discreetness and convenience of vaping.” Viewing this information may prompt those who have used nicotine vapes to transition to (or use dually) cannabis/DICP vapes due to the shared vaping experience. We also found specific instructions for vape device use, which may increase the perceived easiness of product use for young individuals who are new to vaping devices (31). Overall, viewing information about vaping experience and use instructions may reduce the perceived barriers to DICVP use, inviting new consumers to experiment with products.

We also found a large proportion of marketing features related to regulation and compliance, which described that the products were lab-tested for additives and chemicals, the health warnings (all from California and specific for Delta-9 THC products), and the legality of the products under the 2018 Farm Bill. However, it may be challenging for consumers to interpret and verify the provided information, including whether these products were actually lab-tested and whether these labs are accredited or not. Cautions were also raised related to the increasingly falsified lab testing results used by the cannabis industry (32). In addition, viewing information stating the products are lab-tested and legal under the Farm Bill may increase the perceived legality and safety of these products, which may promote use, especially among those in states where traditional cannabis is illegal (6,33). Warnings related to the health risks of cannabis use – all from California Prop 65 and specific to Delta-9 THC – were only found in the descriptions of about half of the products. Evidence has shown that warning labels on cannabis products communicate risks to consumers and have the potential to educate consumers about cannabis harm (34). State laws like the ones in California may help consumers make informed decisions about the risks of product use. However, the fact that these warning messages are specific to Delta-9 THC may reduce the effectiveness of the messages given the diversity of cannabinoids documented in DICPs. This may also lead people to believe these other intoxicating cannabinoids are safer than Delta-9 THC. More research on the health consequences of DICP use is needed to inform warning and preventive messaging.

Flavor and sensation claims, a commonly found marketing strategy in this study, may enhance consumer preferences and curiosity about using DICVPs. Extensive literature on nicotine vape product marketing has shown that flavor-related descriptors (especially fruit flavors), flavor choices, and flavor sensation-related claims are among the most appealing marketing features in their advertisements among young individuals, and that those features often generate an increased willingness to try the products shown in the advertisements (15,35– 37). This may be especially true given artificial flavors from the e-cigarette industry are used for cannabis vapes (38). Although some of the identified flavor names may overlap with product strain names, it conveys youth-friendly taste and aroma that may instigate product curiosity from this group. In addition, we identified a series of trademark names of popular dessert and snack products for describing product flavors, raising concerns that these flavor names may entice young people to try DICVPs. Although local regulations on traditional cannabis and DICVPs have not started restricting product flavors, many jurisdictions prohibit traditional cannabis product marketing that targets minors (39). Studies are needed to examine whether and how DICVP marketing strategies with youth-appealing flavor names and sensation claims may prompt product use.

Promoting psychoactive effects was also a commonly used marketing strategy for DICVPs. Such claims may increase consumers’ intentions to use these products, as achieving a “high” is a widely recognized motivation for cannabis use (40). Other purported positive effects identified—such as mood enhancement, improved socialization, and increased focus or creativity—align with well-established cannabis use expectancies in the literature (41,42). According to the means-end approach in advertising theory, marketing product features highlighting the functions and benefits of product use can shape motives and expectations related to product use, effectively driving interest in use (43). Finally, we found a small number of descriptions stating unfounded or misleading health effect claims (e.g., reducing stress and anxiety) to describe the positive benefits of DICVP use. To prevent deceptive advertising practices, ongoing monitoring of health-related claims for DICVPs – and DICPs overall – is needed (44).

We also found an extensive number of claims indicating product quality, including using descriptors of “natural,” “quality,” “purity,” “premium,” and “organic.” According to the evidence from tobacco marketing literature, some of these descriptors may reduce perceived harm and risks from product use and promote use intentions (45–49). This may also create the “health halo” effect, observed in tobacco and food industry marketing (50), where consumers may overestimate a product’s healthiness based on ingredient claims suggesting the quality or cleanness of the products. Research on how these marketing claims related to high product quality may affect risk perceptions of DICVP use is especially needed because the public has increasingly considered cannabis (and vape products) to be “healthy” and “harm-free” (51,52) — a belief that will become even more common as cannabis products become further normalized.

### Strengths and Limitations

This study used product descriptions from highly visited online retail websites to gather key insights into the existing retail market environment for a large number of DICVPs. However, this study only collected data from two websites, making it unrepresentative of the broader retail market, including brick-and-mortar stores, and it did not examine consumer purchasing habits or product use patterns. Because of the rapidly evolving nature of the DICVP marketplace, continuous monitoring of product attributes and marketing features is needed. Furthermore, our analysis did not examine marketing features shown on product packages, which can also contain youth-appealing features such as cartoons and anime, fruit and candy images, and vibrant colors that may generate product use interest among young people. Lastly, our study did not include cannabis strains as a product attribute due to the high variety of strain names found from the websites. Some strain names “Girl Scout Cookies” and “Gelato” may overlap with product flavor names and be perceived as flavors by the consumers. Since cannabis strains may produce distinct aroma and health effects (53, 54), future studies should further examine the scope of DICVP strains listed on retail websites.

## CONCLUSIONS

This study provides a comprehensive analysis of the current online retail market for DICVPs, highlighting its variety of brands, intoxicating compounds, and marketing features that may appeal to younger or novice consumers. The rapid diversification of this market necessitates continuous monitoring, with future research needed to explore consumer purchasing patterns and use behaviors. Such data are essential for guiding policy, public health practices, and further research efforts related to DICVP marketing. Future research aimed at examining the behavioral effects of these marketing features is greatly needed to understand the most impactful marketing tactics to inform policymaking for reducing product youth appeal.

## Data Availability

All data produced are available on the websites

## REFERENCES

1. Abernethy A. Hemp Production and the 2018 Farm Bill. U.S. Food and Drug Administration; 2019 Jul. Available from: https://www.agriculture.senate.gov/imo/media/doc/Testimony_Abernethy%2007.25.19.pdf

2. Rossheim ME, Tillett KK, Vasilev V, LoParco CR, Berg CJ, Trangenstein PJ, et al. Types and Brands of Derived Psychoactive Cannabis Products: An Online Retail Assessment, 2023. Cannabis and Cannabinoid Research. 2024 Jan 19;can.2023.0266.

3. LoParco CR, Rossheim ME, Walters ST, Zhou Z, Olsson S, Sussman SY. DeltaC8 tetrahydrocannabinol: a scoping review and commentary. Addiction. 2023 Jun;118(6):1011– 28.

4. Rossheim ME, LoParco CR, Walker A, Livingston MD, Trangenstein PJ, Olsson S, et al. Delta-8 THC Retail Availability, Price, and Minimum Purchase Age. Cannabis and Cannabinoid Research. 2024 Feb 1;9(1):363–70.

5. Babalonis S, Raup-Konsavage WM, Akpunonu PD, Balla A, Vrana KE. Δ8-THC: legal status, widespread availability, and safety concerns. Cannabis and Cannabinoid Research. 2021;6(5):362–5.

6. Rossheim ME, LoParco CR, Tillett KK, Treffers RD, Livingston MD, Berg CJ. Intoxicating cannabis products in vape shops: United States, 2023. American Journal of Preventive Medicine. 2024;67(5):776–84.

7. LoParco CR, Henry D, Prater Z, Bone C, Rossheim ME, Berg CJ. “Non-Flavor” Flavors: What Are the Implications of Derived Psychoactive Cannabis Product Marketing? Journal of Studies on Alcohol and Drugs. 2024 Sep;85(5):756–8.

8. Rossheim ME, LoParco CR, Berg CJ, Tillett KK, Trangenstein PJ, Henry D, et al. Derived psychoactive cannabis products and 4/20 specials: An assessment of popular brands and retail price discounts in Fort Worth, Texas, 2023. Drug and Alcohol Dependence. 2024;256:111119.

9. Harlow AF, Miech RA, Leventhal AM. Adolescent Δ8-THC and Marijuana Use in the US. JAMA. 2024;331(10):861–5.

10. Rossheim ME, Loparco CR, Henry D, Trangenstein PJ, Walters ST. Delta-8, Delta-10, HHC, THC-O, THCP, and THCV: What Should We Call These Products? Journal of Studies on Alcohol and Drugs. 2023 May;84(3):357–60.

11. National Academies of Sciences, Engineering, and Medicine, Health and Medicine Division, Board on Population Health and Public Health Practice, Committee on the Health Effects of Marijuana: An Evidence Review and Research Agenda. The Health Effects of Cannabis and Cannabinoids: The Current State of Evidence and Recommendations for Research. Washington (DC): National Academies Press (US); 2017. (The National Academies Collection: Reports funded by National Institutes of Health).

12. LoParco CR, Tillett KK, ChenCSankey J, Berg CJ, Rossheim ME. Public health considerations about tetrahydrocannabinolCinfused beverages. Addiction. 2024 Sep 26;add.16676.

13. Luc MH, Tsang SW, Thrul J, Kennedy RD, Moran MB. Content analysis of online product descriptions from cannabis retailers in six US states. International Journal of Drug Policy. 2020;75:102593.

14. Hinckley JD, Hopfer C. Marijuana legalization in Colorado: increasing potency, changing risk perceptions, and emerging public health concerns for youth. Adolescent Psychiatry (Hilversum, Netherlands). 2021;11(2):95.

15. Chen-Sankey J, Jeong M, Wackowski OA, Unger JB, Niederdeppe J, Bernat E, et al. Noticing People, Discounts, and Non-Tobacco Flavors in E-cigarette Ads May Increase E-cigarette Product Appeal among Non-Tobacco-Using Young Adults. Published online first. Tobacco Control. 2022 Jun 7.

16. Chen-Sankey JC, Kong G, Choi K. Perceived ease of flavored e-cigarette use and e-cigarette use progression among youth never tobacco users. PloS one. 2019;14(2).

17. Chen JC, Green K, Fryer C, Borzekowski D. Perceptions about e-cigarette flavors: a qualitative investigation of young adult cigarette smokers who use e-cigarettes. Addiction Research & Theory. 2019;27(5):420–8.

18. Hasin DS, Borodovsky J, Shmulewitz D, Walsh C, Livne O, Struble CA, et al. Use of highly-potent cannabis concentrate products: More common in U.S. states with recreational or medical cannabis laws. Drug and Alcohol Dependence. 2021 Dec 1;229:109159.

19. Spindle TR, Cone EJ, Schlienz NJ, Mitchell JM, Bigelow GE, Flegel R, et al. Acute effects of smoked and vaporized cannabis in healthy adults who infrequently use cannabis: A crossover trial. JAMA Netw Open. 2018 Nov 30;1(7):e184841.

20. Spindle TR, Martin EL, Grabenauer M, Woodward T, Milburn MA, Vandrey R. Assessment of cognitive and psychomotor impairment, subjective effects, and blood THC concentrations following acute administration of oral and vaporized cannabis. Journal of Psychopharmacology. 2021 Jul;35(7):786–803.

21. Shi Y, Cao Y, Shang C, Pacula RL. The impacts of potency, warning messages, and price on preferences for Cannabis flower products. International Journal of Drug Policy. 2019;74:1– 10.

22. Zhu B, Guo H, Cao Y, An R, Shi Y. Perceived importance of factors in cannabis purchase decisions: a best-worst scaling experiment. International Journal of Drug Policy. 2021;91:102793.

23. Chen-Sankey J, La Capria K, Meng S, Mazzeo R, Vijayakumar N, Padon AA, et al. Product Features Used to Promote Top-selling Cannabis Vape Products in An Online Retail Environment. medRxiv. 2024;2024–11.

24. LoParco CR, Tillett KK, Berg CJ, Rossheim ME. Online retail of derived psychoactive cannabis products: age and shipping restrictions. Journal of Adolescent Health. 2024. 75(2):249–253.

25. Egan KL, Villani S, Soule EK. Absence of age verification for online purchases of Cannabidiol and Delta-8: Implications for youth access. Journal of Adolescent Health. 2023;73(1):195–7.

26. Cavazos-Rehg PA, Krauss MJ, Cahn E, Lee KE, Ferguson E, Rajbhandari B, et al. Marijuana promotion online: An investigation of dispensary practices. Prevention Science. 2019;20:280–90.

27. Kowitt SD, Yockey RA, Lee JG, Jarman KL, Gourdet CK, Ranney LM. The impact of cannabis packaging characteristics on perceptions and intentions. American journal of preventive medicine. 2022;63(5):751–9.

28. Krippendorff K. Measuring the reliability of qualitative text analysis data. Quality and Quantity. 2004;38:787–800.

29. Leas EC, Nobles AL, Shi Y, Hendrickson E. Public interest in? 8-Tetrahydrocannabinol (delta-8-THC) increased in US states that restricted? 9-Tetrahydrocannabinol (delta-9-THC) use. International Journal of Drug Policy. 2022;101:103557.

30. Chan-Hosokawa A, Nguyen L, Lattanzio N, Adams WR. Emergence of delta-8 tetrahydrocannabinol in DUID investigation casework: method development, validation and application. Journal of Analytical Toxicology. 2022;46(1):1–9.

31. Farrelly MC, Duke JC, Crankshaw EC, Eggers ME, Lee YO, Nonnemaker JM, et al. A randomized trial of the effect of e-cigarette TV advertisements on intentions to use e-cigarettes. American Journal of Preventive Medicine. 2015;49(5):686–93.

32. Chemical & Engineering News [cited 2025 Jan 17]. Shopping around for favorable cannabis testing labs. Available from: https://cen.acs.org/biological-chemistry/natural-products/Shopping-around-favorable-cannabis-testing/102/i28

33. Livingston MD, Walker A, Cannell MB, Rossheim ME. Popularity of Delta-8 THC on the Internet Across US States, 2021. American Journal of Public Health. 2022 Feb;112(2):296– 9.

34. Pepper JK, Lee YO, Eggers ME, Allen JA, Thompson J, Nonnemaker JM. Perceptions of US and Canadian cannabis package warnings among US adults. Drug and Alcohol Dependence. 2020;217:108275.

35. Erinoso O, Smith KC, Iacobelli M, Saraf S, Welding K, Cohen JE. Global review of tobacco product flavour policies. Tobacco Control. 2021;30(4):373–9.

36. Chen-Sankey J, Weiger C, La Capria K. Using Eye Tracking to Examine Young Adults’ Visual Attention to E-cigarette Advertising Features and Associated Positive E-cigarette Perceptions. Annals of Behavioral Medicine. 2024 May 23;58(6):445–456.

37. Chen-Sankey J, Weiger C, La Capria K, Vassey J, Jeong M, Phan L, et al. Young Adults’ Visual Attention to Features of Social Media Marketing for Disposable E-cigarettes and Associated Perceptions. Addiction. doi: 10.1111/add.16586. Online ahead of print.

38. What’s In Your Vape Cartridge? Cannabis Business Time. Retrieved from: https://www.cannabisbusinesstimes.com/business-issues-benchmarks/cannabis-oil-product-manufacturing/article/15695031/whats-in-your-vape-cartridge#:~:text=Artificial%20flavors%3A%20Typically%2C%20the%20artificial,from%20the%20e%2Dcigarette%20industry. Retrieved on: January 22, 2025

39. State Regulation of Adult Use Cannabis Advertising [cited 2023 Dec 13]. Available from: https://www.networkforphl.org/wp-content/uploads/2022/11/State-Regulation-of-Adult-Use-Cannabis-Advertising.pdf

40. Bray BC, Berglund PA, Evans-Polce RJ, Patrick ME. A Latent Transition Analysis of Self-Reported Reasons for Marijuana Use During Young Adulthood. Evaluation & the Health Professions. 2021 Mar;44(1):9–24.

41. Lee CM, Neighbors C, Woods BA. Marijuana motives: young adults’ reasons for using marijuana. Addictive Behaviors. 2007 Jul;32(7):1384–94.

42. Buckner JD, Ecker AH, Welch KD. Psychometric properties of a valuations scale for the Marijuana Effect Expectancies Questionnaire. Addictive Behaviors. 2013 Mar;38(3):1629– 34.

43. Reynolds TJ, Olson JC. Understanding consumer decision making: The means-end approach to marketing and advertising strategy. Psychology Press; 2001.

44. Federal Trade Commission. 2021 [cited 2024 Dec 18]. FTC and CBD: Latest case challenges unproven health claims. Available from: https://guidance/blog/2021/05/ftc-cbd-latest-case-challenges-unproven-health-claims

45. Moran MB, Heley K, Czaplicki L, Weiger C, Strong D, Pierce J. Tobacco Advertising Features That May Contribute to Product Appeal Among US Adolescents and Young Adults. Nicotine & Tobacco Research. 2021 Aug 4;23(8):1373–81.

46. Iles IA, Pearson JL, Lindblom E, Moran MB. “Tobacco and Water”: Testing the Health Halo Effect of Natural American Spirit Cigarette Ads and Its Relationship with Perceived Absolute Harm and Use Intentions. Health Communication. 2020 Jan 10;36(7):804–15.

47. Berg CJ, Duan Z, Wang Y, Thrasher JF, Abroms LC, Khayat A, et al. Impact of different health warning label and reduced exposure messages in IQOS ads on perceptions among US and Israeli adults. Preventive Medicine Reports. 2023 Apr 15;33:102209.

48. Duan Z, Levine H, Bar-Zeev Y, Cui Y, LoParco CR, Wang Y, et al. Health warning labels on heated tobacco products and their impact on use intentions and risk perceptions: a cross-sectional study of adult tobacco users in the US and Israel. Israel Journal of Health Policy Research. 2023 Nov 13;12(1):33.

49. Chen-Sankey J, Ganz O, Seidenberg A, Choi K. Effect of a “tobacco-free nicotine” claim on intentions and perceptions of Puff Bar e-cigarette use among non-tobacco-using young adults. Tobacco Control. 2023;32(4):501–4.

50. Williams P. Consumer Understanding and Use of Health Claims for Foods. Nutrition Reviews. 2005 Jul;63(7):256–64.

51. Levy NS, Mauro PM, Mauro CM, Segura LE, Martins SS. Joint perceptions of the risk and availability of cannabis in the United States, 2002-2018. Drug and Alcohol Dependence. 2021 Sep;226:108873.

52. Pacek LR, Mauro PM, Martins SS. Perceived risk of regular cannabis use in the United States from 2002 to 2012: differences by sex, age, and race/ethnicity. Drug and Alcohol Dependence. 2015 Apr;149:232–44.

53. Gilbert AN, DiVerdi JA. Consumer perceptions of strain differences in Cannabis aroma. PLoS One. 2018 Feb 5;13(2):e0192247.

54. Szejko N, Becher E, Heimann F, Grotenhermen F, Müller-Vahl KR. Medicinal use of different cannabis strains: Results from a large prospective survey in Germany. Pharmacopsychiatry. 2024 May;57(03):133–40.

